# A synonymous *SLC2A1* variant causes familial epilepsy and paroxysmal exercise-induced dyskinesia by creating aberrant mosaic splicing patterns

**DOI:** 10.1101/2025.04.05.25325007

**Authors:** Adam T Higgins, Fiona Wang, William O Pickrell, Ann J Johnston, Jeya Natarajan, Khalid Hamandi, Johann te Water Naudé, Mary Chebib, Mark I Rees, Phil E Smith, Seo-Kyung Chung

**Affiliations:** Neurology Research Group, Institute of Life Science, Swansea University Medical School, Swansea University, Swansea, UK; Kids Neuroscience Centre, Kids Research, Children’s Hospital at Westmead, Sydney, Australia; Brain and Mind Centre, School of Medical Sciences, Faculty of Medicine & Health, University of Sydney, Australia; Neurology Department, Morriston Hospital, Swansea Bay University Health Board, Swansea, UK; Neurology Department, University Hospital of Wales, Cardiff, UK; Department of Paediatrics, Royal Glamorgan Hospital, Llantrisant, UK; Paediatric Neurology, University Hospital of Wales, Cardiff, UK

**Author notes:** Corresponding Author: Associate Professor Seo-Kyung Chung, Brain & Mind Centre, Faculty of Medicine & Health, University of Sydney, Australia.

**Keywords:** Glucose Transporter type-1, Deficiency syndrome, VUS, Gene-Splicing, Mosaicism, Families

## Abstract

**Background:** Glucose transporter type-1 deficiency syndrome (GLUT1-DS) arises from variants in the *SLC2A1* gene encoding the glucose transporter type-1 (GLUT1). Genetic analysis of a GLUT1-DS family identified a recurrent heterozygous synonymous *SLC2A1* variant, adjacent to a 5’ donor splice site (NG_008232.1(NM_006516.4): c.972G>A, NP_006507.2: p.Ser324=). The splice site proximity and family segregation analysis warranted an investigation into *SLC2A1* mRNA splicing. The same genotype has been published in two further GLUT1-DS multiplex families without functional biology validation.

**Methods:** The proband exhibited juvenile onset focal epilepsy and paroxysmal exercise induced dyskinesia and family members underwent multiplex segregation analysis for c.972G>A. Family members exhibited phenotypes including focal epilepsy, intellectual disability, early-onset absence epilepsy and paroxysmal exercise-induced dyskinesia. *In silico* and *in vitro* minigene analysis assessed the effect of c.972G>A on *SLC2A1* mRNA splicing.

**Results:** Segregation analysis revealed the synonymous variant associates with more severe epilepsy and GLUT1-DS phenotypes in the family. *In silico* analysis predicted a disruption of the 5’ donor splice site of intron 7. *In vitro* minigene assays demonstrated the activation of two cryptic donor splice sites, generating three transcripts: WT, and two aberrantly spliced isoforms causing 4bp and 32bp deletions. This was previously undetectable by whole blood RNA analysis.

**Conclusions:** Our report demonstrates the synonymous c.972G>A, p.Ser324= variant causes a leaky mosaic *SLC2A1* splicing aberrancy, affecting ∼40% of transcripts, with ∼60% remaining WT spliced. These GLUT1 frameshift deletions result in a variable GLUT1 haploinsufficiency and phenotypic heterogeneity in 3 GLUT1-DS families and highlights the clinical importance of synonymous variants.

## INTRODUCTION

As an important determinant of energy metabolism, the glucose transporter type-1 (GLUT1) governs the availability of glucose in the human brain, encoded by the solute carrier family 2, member 1 gene (*SLC2A1;* 1p34.2; MIM 138140) [1]. GLUT1 is a class-1 member of the GLUT family comprised of 14 transmembrane proteins that facilitate the transport of monosaccharides, polyols, and other compounds across the eukaryotic plasma membrane [2, 3]. Comprising 492 amino acids, GLUT1 functions as the principal transporter of glucose across the blood-brain barrier (BBB) and astrocyte plasma membrane [4]. Consequently, any genetic defect affecting GLUT1 can impair the transportation of glucose across the BBB and into astrocytes, precipitating a cerebral energy deficit termed glucose transporter type-1 deficiency syndrome (GLUT1-DS; OMIM: 606777). Homozygous *SLC2A1* loss is embryonic lethal whilst heterozygous haploinsufficient *SLC2A1* mutations result in a marked paucity of cerebral glucose levels, severe depletion of brain energy reserves and neuroglycopenia [5]. The culmination is embodied in the infantile-onset neurodevelopmental disorder, GLUT1-DS [6].

GLUT1-DS is a rare genetic neurometabolic disease consisting of multiple overlapping phenotypes, including generalised epilepsy, dystonic tremor, paroxysmal exercise induced dyskinesia (PED) and intellectual disability (ID) [6–12]. More than 200 genetic variants have been reported in GLUT1-DS patients and families, including missense, nonsense, splicing, frameshift and in-frame insertions/deletions and microdeletions [8, 9, 13, 14]. Previous studies have categorised GLUT1 variants by their levels of neurological impairment using the semi-quantitative Columbia Neurological Score [15]. Missense variants predominantly occurred in those with mild to moderate neurological impairment. Splice site, nonsense and insertions / deletions occurred mainly in the moderate to severe categories, with complete gene deletions grouped in the severe category [4]. Current treatment for GLUT1-DS includes a ketogenic (high-fat, low-carbohydrate) diet, effective in reducing seizure frequency and dyskinesias [16].

While missense, nonsense, and insertion/deletion mutations have been extensively studied in GLUT1-DS, the contribution of splicing alterations remains underexplored. Among the 200+ GLUT1-DS-associated genetic variants reported in the Human Gene Mutation Database (HGMD; **supplemental figure 1**), only 22 variants are identified as splicing mutations, including 4 intronic variants and 18 essential splice site variants (HGMD database, January 2024). However, the precise impact of these variants on GLUT1 protein expression and function is not fully characterised.

In our study of a large multiply-affected GLUT1-DS family, we identified a *SLC2A1* synonymous variant, NG_008232.1(NM_006516.4):c.972G>A, NP_006507.2:p.Ser324=. This variant has been previously reported in two other multiplex families with paroxysmal exercise-induced dyskinesia, however, the analysis of whole blood RNA did not detect *SLC2A1* mRNA splicing aberrancies [17, 18]. The location of this synonymous variant adjacent to exon-intron boundaries suggested the possibility of altered GLUT1 splicing patterns and has a VUS status of potentially pathogenic. With updated platforms and emerging evidence of synonymous splicing aberrancy in genetic disease [19], our study conducted further analysis of *SLC2A1* c.972G>A, using a minigene splicing system, resulting in clinically-impactful findings.

## METHODS

### Genetic Analysis

Sequencing was commissioned on proband DNA using an epilepsy gene panel service based at the Glasgow Children’s Hospital and submitted as part of the Epi4K consortium epilepsy families project [20]. The diagnostic service identified a heterozygous c.972G>A variant in the *SLC2A1* gene, and seven family members underwent segregation analysis by Sanger sequencing. The genomic region encompassing *SLC2A1* exons 7 & 8 (with flanking introns) was PCR amplified with the Multiplex PCR Kit (QIAGEN, Cat: 206143) and purified with the QIAquick PCR Purification kit (QIAGEN, Cat: 28106), according to the manufacturer’s protocols. Primers used were: forward (5’-AGTGTCCCTTCTGCCTGAGTA-3’), reverse (5’-CAGGCATTTTGGGATATGAAGCC-3’). Purified DNA products were Sanger sequenced (Eurofins, MWG Operon, Germany) and aligned in Mutation Surveyor V4.20 (SoftGenetics) against the reference *SLC2A1* sequence (NCBI, NG_008232.1). HGVS 21.0.2 variant nomenclature guidelines were used in this report.

### Bioinformatic Analysis

The c.972G>A *SLC2A1* variant was analysed using *in silico* mRNA splicing tools to predict potential splicing outcomes. Eight tools were used: Human Splice Finder (HSF 3.1), NetGene2, NNSPLICE, Fsplice, MaxENTScan [10], SpliceAI, SROOGLE and CRYP-SKIP. All programs provide scores for changes in the wild-type (WT) splice site strength and alterations in cryptic splice site strengths in the presence of a nucleotide variant. Details on scoring and thresholds for the programs are outlined in **supplemental table 1**.

### Minigene Splicing Assay

An *in vitro* splice assay utilising the pET01 Exontrap system (MoBiTech, Cat: K2010) was conducted to determine the *SLC2A1* mRNA splice outcome of the c.972G>A variant (**supplemental Fig. 2**). Minigene splice constructs were generated by inserting a defined genomic region (exons and introns) into the pET01 vector. Constructs were transfected into HEK293 cells for RNA extraction, RT-PCR and PCR analysis. The pET01 vector was chosen due to its intrinsic splicing activity, with two exons flanking the Multiple Cloning Site (MCS).

Primers were designed to capture and amplify the genomic region encompassing exons 5 to 8 of *SLC2A1* with >500bp flanking intron (2,289bp total) in the proband (IV-2), mother (III-1), father (III-2) and sibling (IV-3) with long-range PCR (Q5 HF HotStart Polymerase, NEB). The primers incorporated *XhoI/SacII* restriction digest cut sites (underlined, nucleotide changes in red) corresponding to the pET01 vector MCS: forward primer (5’-CATGGCCTCGAGGTTGACAGCAAGATGAC-3’), reverse primer (5’-GGCTGCCGCGGA GTAAGCGCTAAGCATA-3’). Following purification, the amplicon and vector were separately subject to restriction digestion to generate complementary overhangs. Ligation of the digested amplicon and vector was facilitated with the T4 ligase system (NEB), with the ligated plasmid heat-shock transformed into chemically competent DH5α *E.coli* and plated on Ampicillin (100µg/mL) agar plates. Colonies were picked for PCR to verify the successful insertion of the amplicon into the pET01 vector. Selected colonies were grown in LB broth for subsequent plasmid purification with the QuickLyse MiniPrep Kit (QIAGEN). Purified plasmids were outsourced for Sanger sequencing to determine variant status (WT or c.972G>A from the proband and mother, WT only from father and sibling). Minigene plasmids were classified by the genomic insertion and variant status (i.e. pET01-*SLC2A1*-ex5_8-WT or c.972G>A).

WT and c.972G>A plasmids were transfected into seeded HEK293 subcultures using Turbofectin^TM^ (OriGene). Forty-eight hours post-transfection, cells were lysed, and total cell RNA was extracted with the RNeasy Plus Mini kit (QIAGE), following the manufacturer’s protocol. RNA was subjected to reverse-transcriptase PCR for cDNA synthesis (ImProm-II™ Reverse Transcription System, Promega). cDNA was PCR amplified with the Multiplex PCR Kit and primers targeting the pET01 endogenous exons flanking the inserted genomic exons. PCR products were purified, and agarose gel was electrophoresed to visualise any size difference between WT and c.972G>A spliced products and validated by Sanger sequencing.

### Blue-white Screening of Minigene Spliced Amplicon

Blue-white screening was conducted to isolate and quantify differentially spliced transcripts from the proband minigene assay PCR sample. The purified PCR amplicon from the minigene splicing assay was ligated with the pGEM®-T Easy Vector (Promega) using T4 Ligase (Promega) and heat-shock transformed into chemically competent JM109 *E.coli* (Promega). The *E.coli* was plated on IPTG/X-gal supplemented agar plates and white colonies, indicating successful ligation, were picked for PCR amplification of the ligated insert using the Multiplex PCR Kit. Amplicons were subjected to sequential digestion with *AleI* (NEB) and *HincII* (NEB) restriction enzymes to identify WT, 4bp deletion and 32bp deletion amplicons. A selection was purified and Sanger sequencing analysed. Data was graphed using GraphPad Prism (10.2.2).

## RESULTS

Gene panel sequencing and Sanger segregation analysis identified a *SLC2A1* c.972G>A variant in a multiple-affected family presenting with a complex GLUT1-DS phenotype. *In silico* analysis predicted potential splicing alterations downstream of the c.972G>A variant, suggesting a whole or partial exon 7 skipping and subsequent mRNA modifications. Minigene splicing assays unveiled three distinctly spliced transcripts, WT and two aberrant partial exon deletions, with the aberrant transcripts likely leading to non-functional GLUT1 protein.

### GLUT1-DS family

The family presents with a complex GLUT1-DS phenotype, including early-onset absence epilepsy, focal epilepsy, paroxysmal exercise-induced dyskinesia, and intellectual difficulties (**Fig. 1a**). The proband (IV-2) exhibited focal aware sensory seizures in late childhood, accompanied by episodes of leg twitching and cramping, along with abnormal exercise-induced dystonic leg posturing. Similar epilepsy phenotypes were observed in the proband’s mother (III-1) and two children (V-1 and V-2), with additional intellectual disabilities noted in one of the children (V-2) and early-onset absence epilepsy in the proband’s sibling (IV-3). Detailed anonymised clinical summaries are presented in **Table 1**, & **Supplemental Material**.

**Fig. 1:**
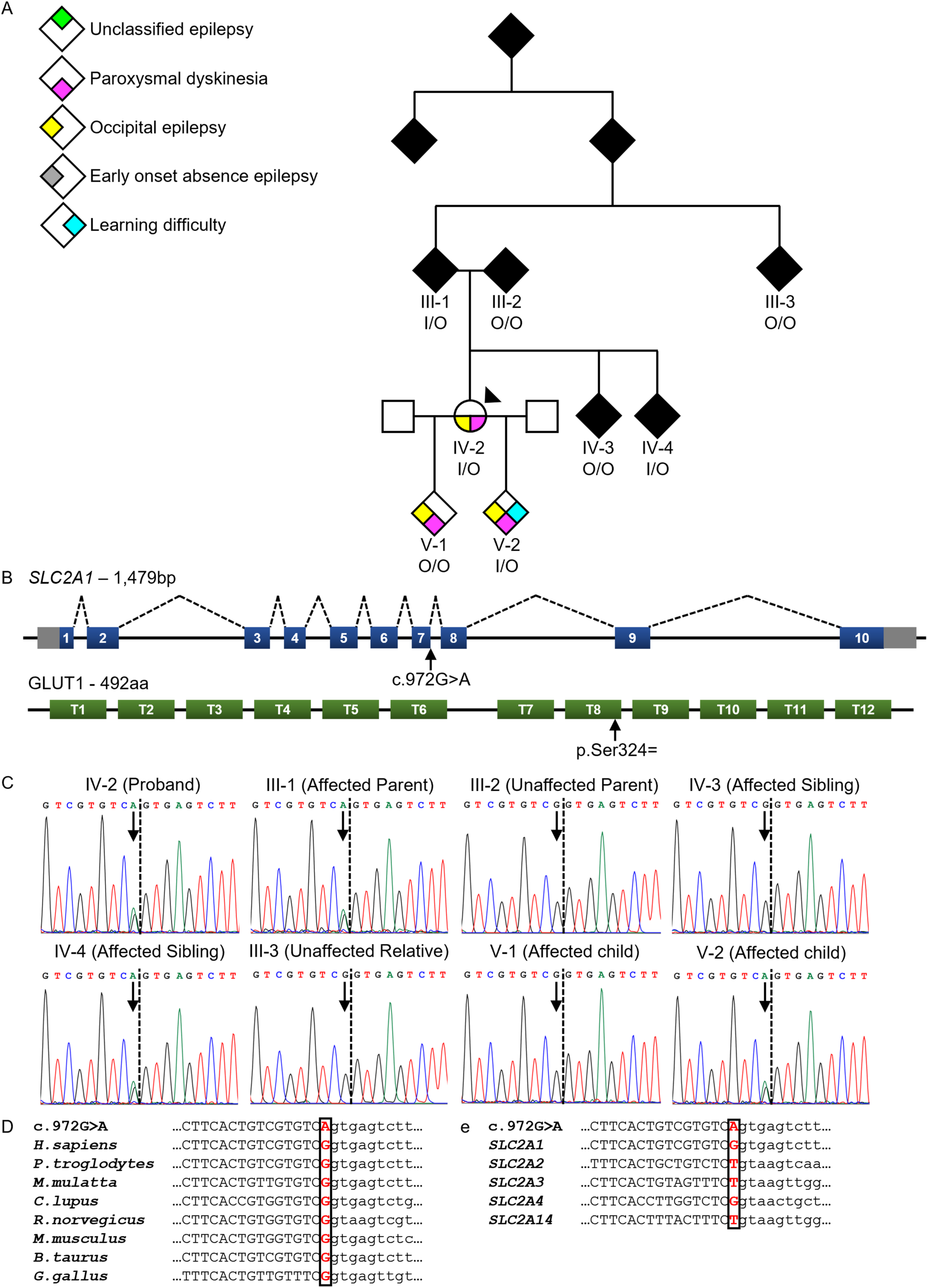
The GLUT-DS family harbouring the *SLC2A1* c.972G>A, p.Ser324= variant. A) The family pedigree with phenotype information shown by colour and the proband (IV-2) arrowed. The c.972G>A variant status of specific family members is denoted with I & O (I=variant, O=WT). Further generations have been anonymised (black diamond shapes) to comply with medRxiv policies, but a full pedigree will be published in the final version. B) A schematic of the *SLC2A1* gene and GLUT1 protein, with the position of the variant and synonymous mutation arrowed. The wild-type *SLC2A1* mRNA splicing pattern is shown by the black dotted lines. T = Transmembrane domain, bp = base pair, aa = amino acids. C) Segregation analysis of the variant shown by Sanger sequencing chromatograms. The exon/intron boundary is shown by the black dotted line with the variant position arrowed. D) The complete conservation of the c.972G nucleotide in mammals. The variant position is shown in bold red font. e) A partial conservation of the G nucleotide in the *SLC2* genes encoding the class-1 GLUT proteins (GLUT1-4, GLUT14). Alignment was performed using Clustal Omega Multiple Sequence Alignment tool.

**Table 1:**
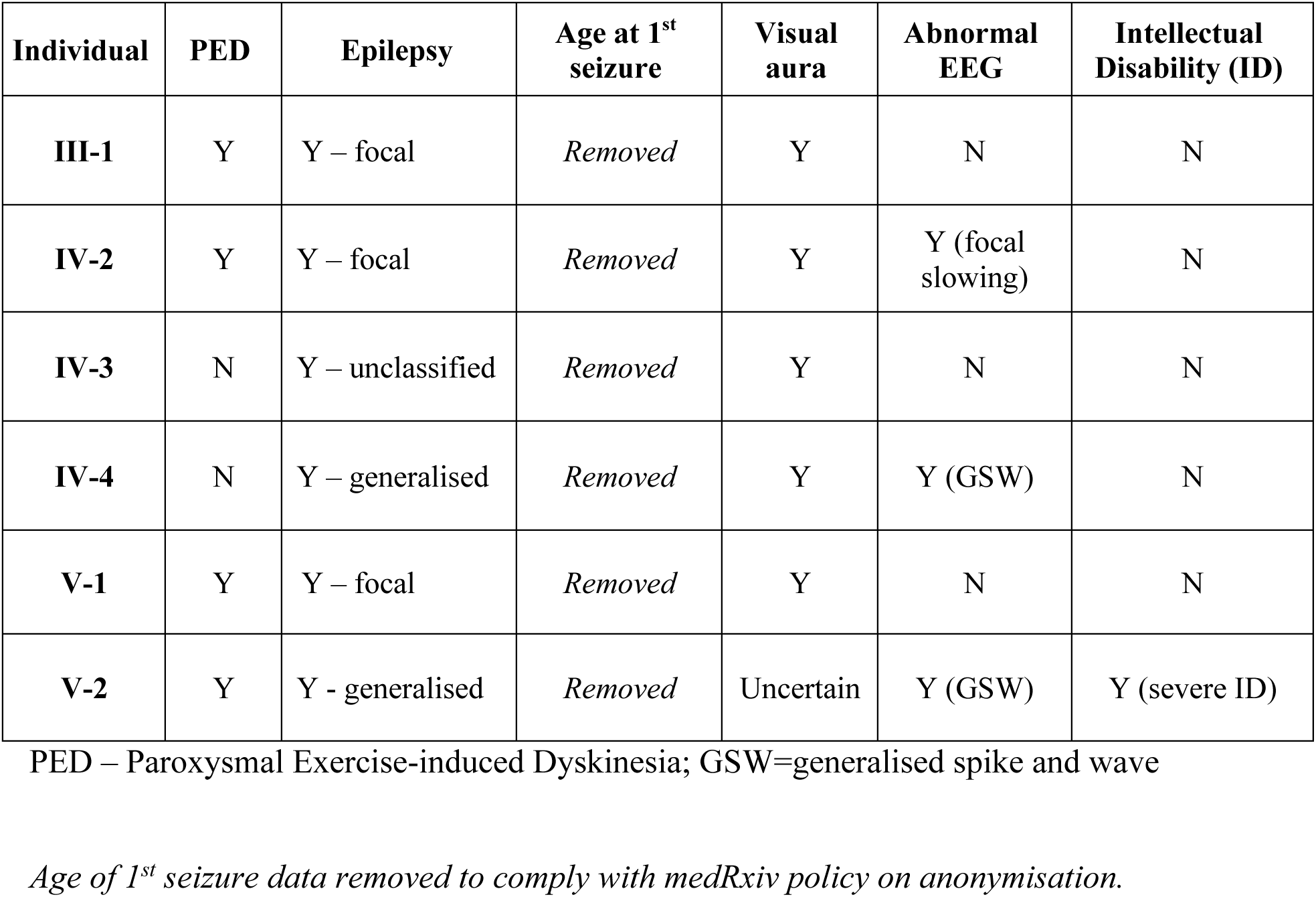
Phenotypic features of the multiplex family presented in Figure 1.

| **Individual** | **PED** | **Epilepsy** | **Age at 1<sup>st</sup> seizure** | **Visual aura** | **Abnormal EEG** | **Intellectual Disability (ID)** |
| --- | --- | --- | --- | --- | --- | --- |
| **III-1** | Y | Y – focal | *Removed* | Y | N | N |
| **IV-2** | Y | Y – focal | *Removed* | Y | Y (focal slowing) | N |
| **IV-3** | N | Y – unclassified | *Removed* | Y | N | N |
| **IV-4** | N | Y – generalised | *Removed* | Y | Y (GSW) | N |
| **V-1** | Y | Y – focal | *Removed* | Y | N | N |
| **V-2** | Y | Y - generalised | *Removed* | Uncertain | Y (GSW) | Y (severe ID) |

PED – Paroxysmal Exercise-induced Dyskinesia; GSW=generalised spike and wave
*Age of 1<sup>st</sup> seizure data removed to comply with medRxiv policy on anonymisation.*

*SLC2A1* candidate-gene screening in the proband identified a heterozygous synonymous variant NG_008232.1(NM_006516.4): c.972G>A, NP_006507.2:p.Ser324=, positioned as the last nucleotide of exon 7 (**Fig. 1b**). Segregation analysis demonstrated an association between the variant and a more severe epilepsy phenotype within the family (**Fig. 1c**). Phylogenetic conservation analysis revealed the mammalian significance of p.Ser324= (**Fig. 1d**). The location, conservation and segregation characteristics prompted further molecular investigation.

### *In silico* splice analysis

Bioinformatic analysis of the c.972G>A *SLC2A1* mRNA splicing variant predicted several potential outcomes (**Fig. 2**). The 5’ essential splice site immediately downstream of the variant position is weakened, and three potential 5’ cryptic donor sites (CDS) identified (two in exon 7, one in intron 7). This is predicted to cause partial exon deletions or intron insertions in the event of a skipped exon 7 from the mRNA transcript.

**Fig. 2:**
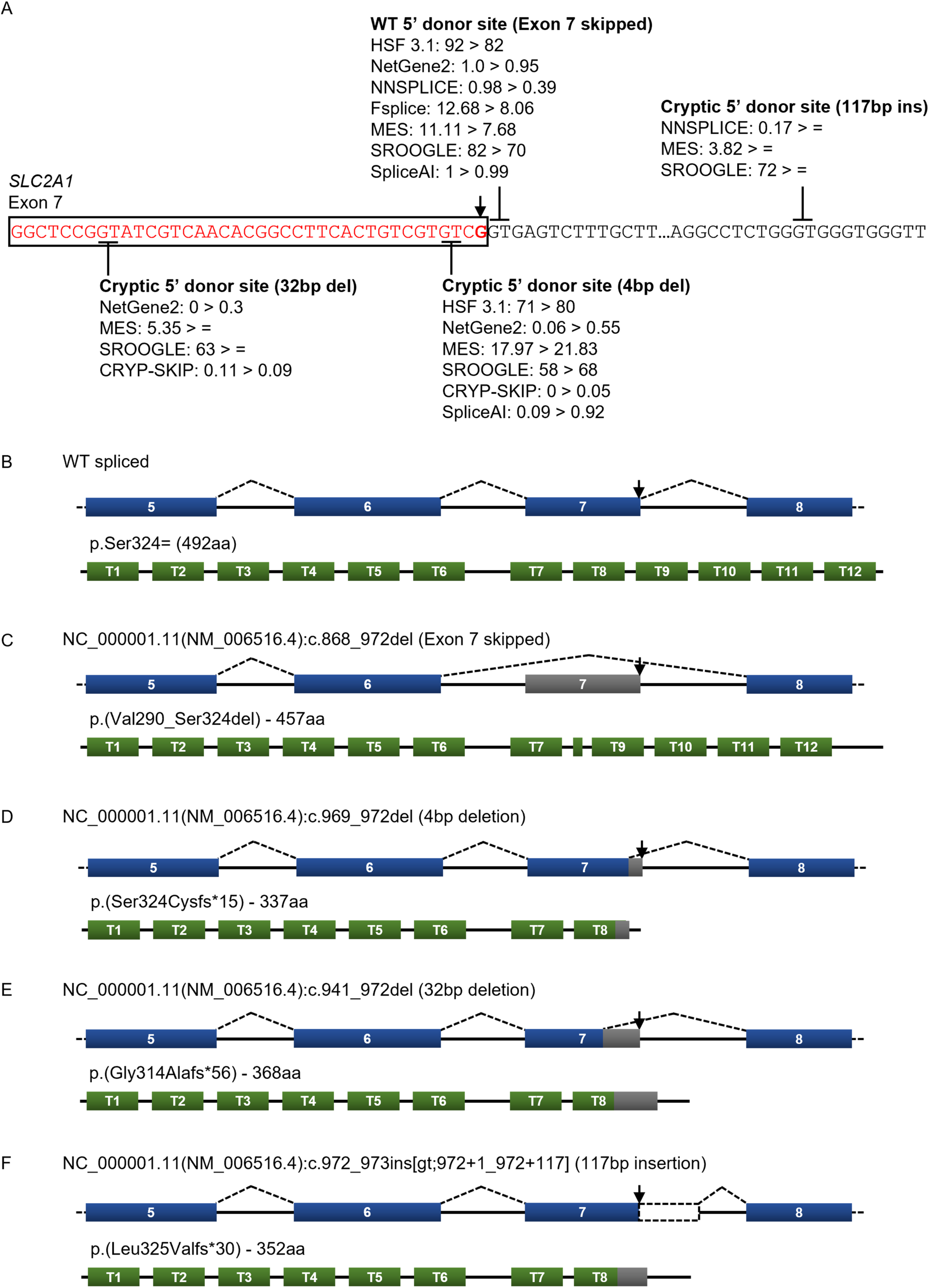
The *in silico* analysis of the *SLC2A1* c.972G>A variant. A) A summary of the *in silico* WT and cryptic splice site (CSS) scoring from several bioinformatic tools for each splice site, in both the WT and variant scenario. The WT splice site in the presence of the variant is weakened in 6 out of 7 tools, with two exonic CSS strengthened (32bp del and 4bp del). A third intronic CSS (117bp ins) remains unaffected by the variant. The positions of the WT and cryptic splice sites are shown with solid black lines. The variant located in the last nucleotide of exon 7 is arrowed. Exon 7 is in red font, intron 7 is in black font. b-f) The predicted spicing pattern of the *SLC2A1* mRNA and predicted consequences on GLUT1 protein in the presence of the variant. Five potential outcomes are predicted: WT spliced harbouring the synonymous variant (B), exon 7 skipping (C), partial deletion of 4bp or 32bp from exon 7 (D and E), partial insertion of 117bp of intron 7 (f). T = Transmembrane domain, bp = base pair, aa = amino acids

Seven out of eight *in silico* programs predicted a reduction in the strength of the relatively strong WT donor splice site in the presence of c.972G>A (**Fig. 2a**). This prediction results in an incomplete exon-skipping event, with transcripts splicing as WT (**Fig. 2b**) and with a deletion of 105bp (NC_000001.11(NM_006516.4):c.868_972del) from the *SLC2A1* mRNA (**Fig. 2c**). Exon 7 skipping causes an in-frame deletion of 35aa from the GLUT1 protein, generating a p.(Val290_Ser324del) outcome.

Two CDS were identified as potential alternative donor sites within exon 7 due to the weakened WT donor site. A CDS at position c.969_970, 4bp upstream of the WT donor site, was identified by six of the seven programs. The use of this CDS predicts a partial exon 7 deletion of 4bp, NC_000001.11(NM_006516.4):c.969_972del (**Fig. 2d**). This deletion was predicted to cause a frameshift deletion from the GLUT1 protein, p.(Ser324Cysfs*15), significantly truncating the protein from 492aa to 337aa. The second CDS was identified 32bp upstream of the WT donor site, at position c.940_941, by four of the seven programs. This CDS predicts a 32bp deletion, NC_000001.11(NM_006516.4):c.941_972del (**Fig. 2e**), truncating the GLUT1 protein from 492aa to 368aa, p.(Gly314Alafs*56). An additional CDS was identified in intron 7 by four of the seven programs, however, no changes in its relative strength were predicted in the presence of the upstream c.972G>A variant. Utilisation of this CDS was predicted to cause an insertion of 117bp of intron 7, NC_000001.11(NM_006516.4):c.972_973ins [gt;972+1_972+117] (**Fig. 2f**), resulting in a frameshift truncation of GLUT1 from 492aa to 352aa, p.(Leu325Valfs*30).

### Minigene splicing assay

The minigene splicing assay using the pET01-*SLC2A1*-ex5_8-WT plasmids derived from proband (IV-2), mother (III-1), father (III-2) and sibling (IV-3) revealed correct *SLC2A1* splicing of exons 5 to 8. The pET01-*SLC2A1*-ex5_8-c.972G>A plasmids derived from the proband and mother revealed a splicing aberrancy, with four distinct transcripts, including WT (**Fig. 3**).

**Fig. 3:**
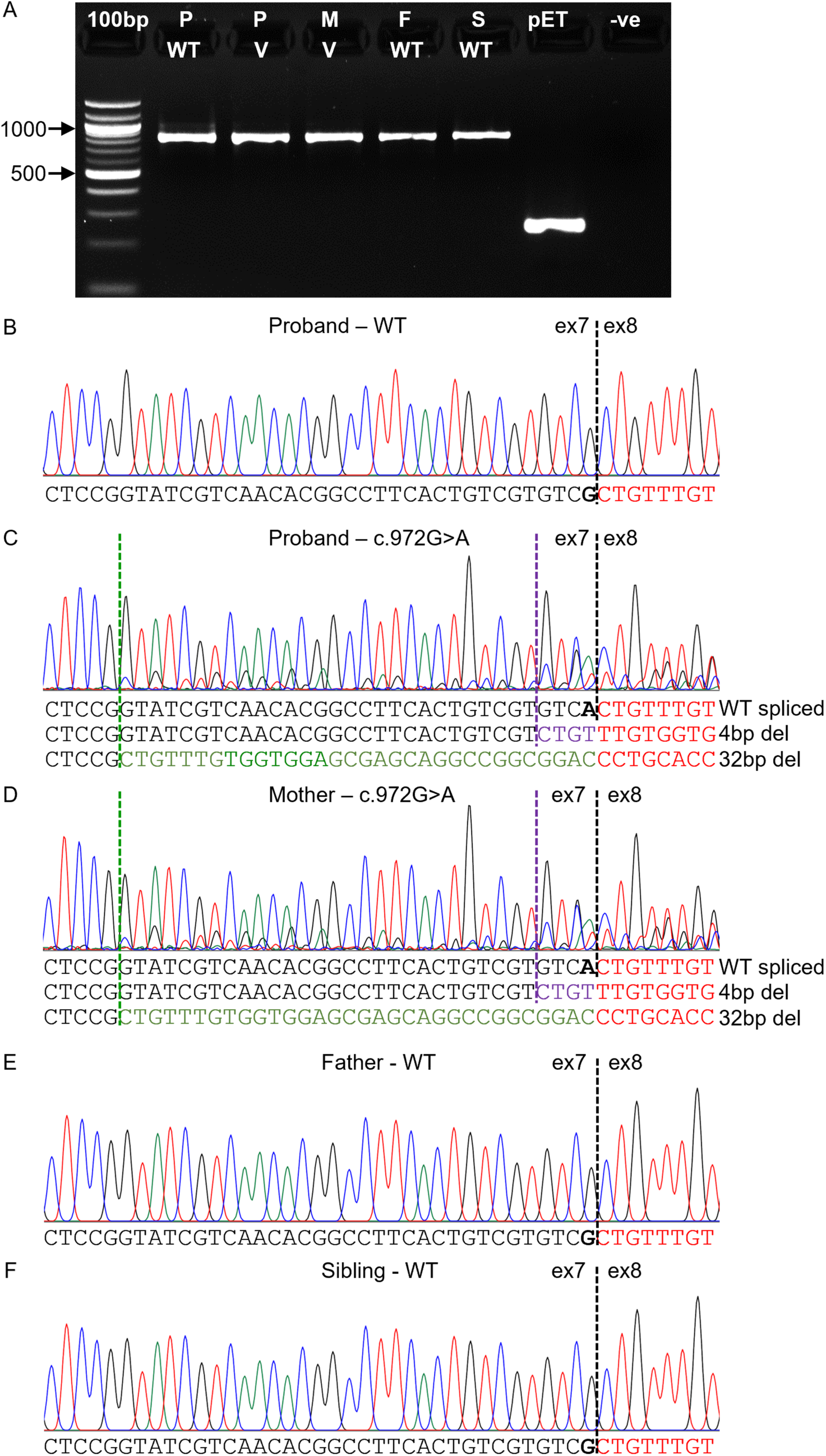
Minigene splicing assay results of *SLC2A1* WT and c.G972A splicing constructs. A) Agarose gel electrophoresis image depicting the cDNA PCR products for WT (800bp), variant (800bp), and pET vector (242bp). cDNA derived from transfected HEK293 cells all appeared to splice as WT (∼800bp). 1000bp and 500bp band on the DNA ladder are arrowed. bp = base pair, P = proband (IV-2), M = Mother (III-1), F = Father (III-2), S = Sibling (IV-3), pET = pET vector, V = variant, -ve = DNA negative PCR control. B-F) Sequence chromatograms of cDNA PCRs for Proband WT and c.G972A, Mother c.G972A, Father WT and Sibling WT. The WT exon 7/exon 8 boundary is indicated by a black dotted line. Sequencing of the proband and mother c.972G>A PCRs showed three transcripts present: a predominant WT spliced transcript carrying the c.972G>A variant, a transcript with 4bp of exon 7 deleted (c.969_972del), and the weakest transcript lacking 32bp of exon 7 (c.941_972del). Exon boundaries due to 4bp deletion (purple dotted line) and 32bp deletion (green dotted line) are highlighted. The variant nucleotide is bolded.

Initial analysis of the minigene cDNA amplicons by agarose gel electrophoresis showed no difference in fragment size between variant and WT plasmids (**Fig. 3a**). WT plasmids were expected to generate an amplicon of 800bp (558bp of *SLC2A1* exons combined with 242bp of pET01 exons). Sanger sequencing of WT amplicons from the proband, father and sibling showed correct splicing of *SLC2A1* exons (**Fig. 3b, e, f**).

Analysis of the Sanger sequencing chromatographs from the amplicons generated by the proband and mother c.972G>A plasmids showed three transcripts (**Fig. 3c-d**): 1) a predominant WT spliced transcript carrying the c.972G>A variant, 2) a transcript with 4bp of exon 7 deleted (c.969_972del) and 3) a transcript lacking 32bp of exon 7 (c.941_972del). Both aberrant transcripts were predicted by *in silico* splicing programs (**Fig. 2d-e**).

The presence of the three transcripts was confirmed with pGEMT-easy blue & white subcloning system. A total of 184 white colonies from proband and 76 from mother were PCR amplified, fractioned, and sequentially digested with *AleI* and *HincII* restriction enzymes to quantify the ratio of the three transcripts. *AleI* digestion cuts WT transcript only, with the 4bp and 32bp transcripts lacking an *AleI* cut site (**Fig. 4a-b**). *HincII* digestion of the remaining amplicons only occurs in the 4bp deletion and WT transcripts due to the 32bp transcript lacking the *HincII* cut site (**Fig. 4a, d**). Sanger sequencing of a selection of amplicons confirmed the presence of WT, 4bp deletion and 32bp deletion transcripts (**Fig. 4b-d**).

**Fig. 4:**
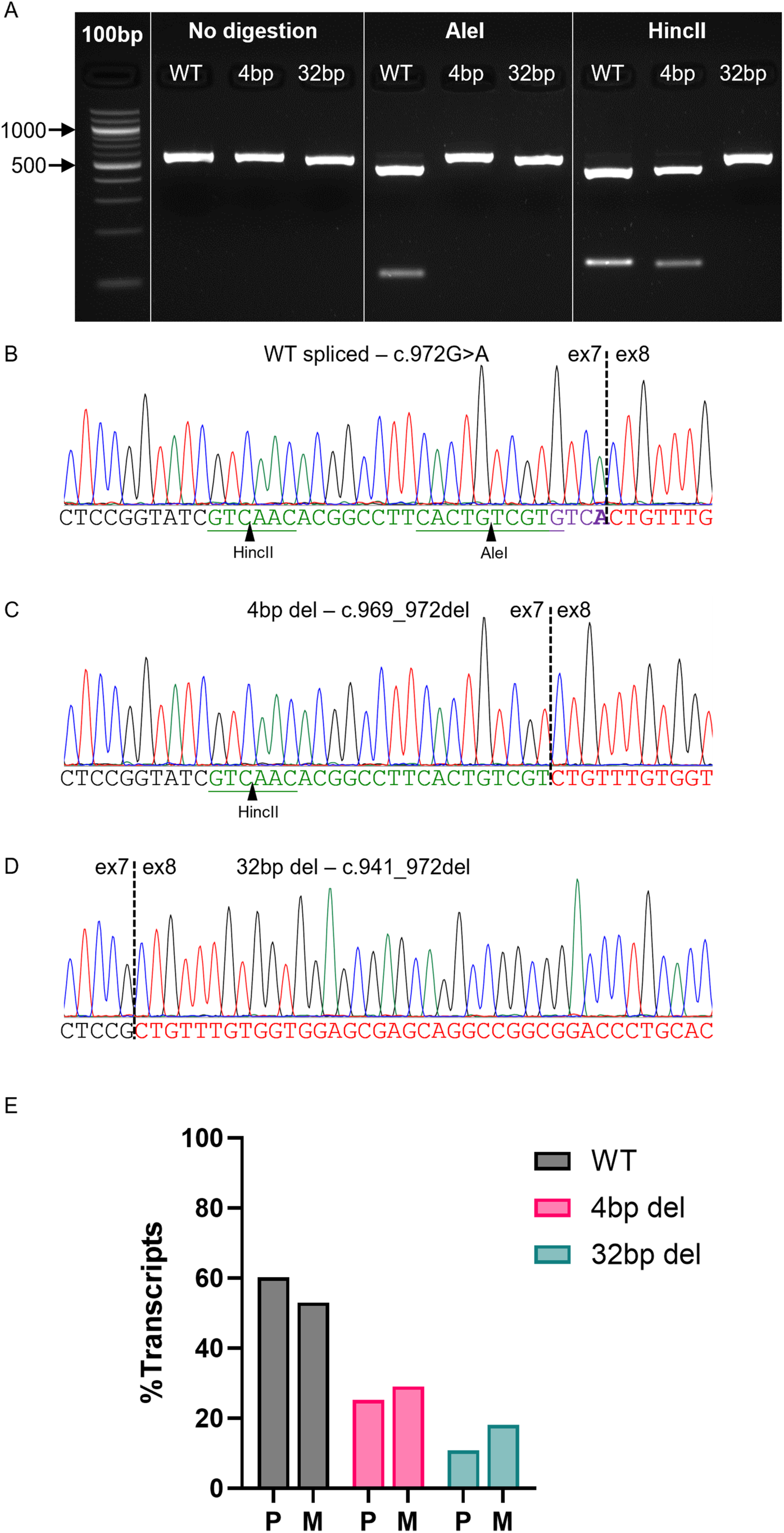
Blue-white colony screening of the Proband (P) and Mother (M) c.G972A cDNA pool. A) Agarose gel with representative PCR products of the three isolated splice isoforms, showing the restriction digestion pattern when digested by *AleI* or *HincII* restriction enzymes. *AleI* cuts only WT transcripts, *HincII* cuts WT and 4bp deleted transcripts.1000bp and 500bp band on the DNA ladder are arrowed. B-D) The sequence chromatograms of the three splice transcripts isolated c.G972A cDNA: WT spliced, 4bp deletion, and 32bp deletion. The WT spliced transcript harbours the variant. The positions of the *AleI* and *HincII* cut sites are underlined and arrowed. E) The proportion of the three transcripts in the c.G972A cDNA pool. Sequential digestion of cDNA PCR products by *AleI* to identify WT transcripts, and *HincII* to identify 4b del transcripts allowed quantification of transcripts in 184 proband PCR products and 76 Mother PCR products. Approximately 60% of proband amplicons digested as WT spliced, 25% as 4bp del, and 15% as 32bp del. Approximately 53% of mother amplicons digested as WT spliced, 29% as 4bp del, and 18% as 32bp del. P = proband, M = mother

Sequential restriction digestion showed that 60% of amplicons from the proband spliced as WT, 25% as 4bp deletion and 15% as 32bp deletion, creating a mosaic expression profile (**Fig. 4e**). These deletions result in frameshift variants in 40% of the spliced transcripts, with 60% remaining WT spliced. The 4bp and 32bp likely leads to GLUT1 truncations, p.(Ser324Cysfs*15) and p.(Ser314Alafs*56), and the loss of functional GLUT1 protein due to mRNA nonsense-mediated decay (NMD). A similar amplicon ratio was observed in the carrier mother. This isoform ratio was reflected in the relative strengths of the individual signals in the splice assay sequencing trace (**Fig. 3d**).

## DISCUSSION

This study investigates the functional consequences of a synonymous *SLC2A1* variant adjacent to the donor splice site in a multi-generational family diagnosed with GLUT1-DS. Genetic screening of *SLC2A1* revealed a c.972G>A variant associated with a more severe phenotype. While previous whole blood RNA analysis showed no splicing aberrancies [17], subsequent and modernised *in silico* and *in vitro* assays uncovered a leaky mosaicism splicing defect linked to the synonymous variant p.Ser324=. This study highlights the pathogenic potential of synonymous variants through splicing aberrations. Further investigation into splice-affecting variants in GLUT1-DS and other epilepsy-related phenotypes and genes could underscore the molecular underpinnings between splicing defects and disease severity. It also shows, across genetic disorders and housekeeping functions, the importance of synonymous variants and their impact on splicing, expression and gene regulation.

The *SLC2A1* p.Ser324= variant was previously published in two independent multiplex families in Germany and Turkey exhibiting familial paroxysmal exercise-induced dyskinesia and absence seizures [17, 18]. Our findings provide pathogenic validation with diagnostic implications for these families. The splicing aberrancy was not detected with whole blood RNA analysis in a previous study, however, this is likely due to the heterozygous nature of the variant and further dilution by the transcript mosaicism [17]. In our minigene splicing, each splicing construct (WT and c.972G>A) mimicked a homozygous WT status with the variant producing a subtle leaky splicing aberrancy. In a heterozygous setting, the aberrancy is likely diluted further, with aberrant splicing possibly dismissed as an artefact or below detection sensitivity. However, due to the partial nature of the splice aberrancy, a mild haploinsufficiency of the GLUT1 protein is likely [15].

Our investigation into the c.972G>A variant’s functional consequences revealed partial or leaky splicing, a phenomenon characterised by incomplete splicing variants associated with low penetrance and reduced expressivity [21]. Such variants can lead to transcript mosaicism, resulting in the production of wild-type full-length reference transcripts alongside aberrantly spliced transcripts [22]. Previous documented cases reporting leaky splicing defects speculate that residual levels of WT protein leads to reduced disease severity or variability in clinical presentation [23]. Whilst transcript mosaicism is likely less common than fully pathogenetic splicing aberrancies, this molecular mechanism has been reported in epilepsy and other neurological disorders [24]. The c.463+5 G>A variant in *CDKL5*, associated with epileptic encephalopathy, weakens the intron 7 donor splice site, causing skipping of exon 7 in ∼50% of *CDKL5* transcripts [25]. Similarly, in the neurofibromatosis type-1 gene (*NF1*), a study of 54 novel variants revealed the c.3362A>G, p.Glu1121Gly missense variant exhibited a mosaic transcriptional profile [26]. Carriers of variants associated with leaky splice mutations tend to exhibit milder phenotypes, suggesting a correlation between leaky splicing and mild haploinsufficiency [27]. To date, eighteen splice site variants have been reported in GLUT1-DS patients (HGMD database [28]). Two studies have reported other splice variants in proximity to the c.972G nucleotide: the c.972+1G>T and the c.972+5G>C variants [29, 30]. The heterozygous c.972+5G>C variant revealed two splicing transcripts, WT and intron 7 retention, creating a GLUT1 truncation frameshift insertion in a proband with moderate intellectual disability, atypical absence seizures and non-convulsive status epilepticus [29]. The c.972+1G>C variant is an essential splice site change likely to result in exon skipping in a GLUT1-DS proband with seizures and intellectual disability [30].

GLUT1-DS individuals with splice variants typically fall into the moderate/severe category for neurological impairment [4]. Previously, 53 GLUT1-DS patients were categorised by their levels of neurological impairment using the semi-quantitative Columbia Neurological Score test (40 to 49 = severe impairment, 50 to 59 = moderate, 60 to 69 = mild, 70 to 76 = minimal) [31] [15]. In that study, missense *SLC2A1* variants predominantly occurred in those with mild to moderate neurological impairment, whereas splice site, nonsense and insertions/deletions occurred mainly in the moderate to severe categories, with complete gene deletions grouped in the severe category [31]. Approximately 90% of GLUT1-DS individuals have *de novo SLC2A1* variants, with the remaining 10% inheriting variants in an autosomal dominant manner [17]. Most individuals with *de novo SLC2A1* mutations exhibited the more severe phenotype, with the familial inherited mutations mostly resulting in a milder GLUT1-DS phenotype [32]. Rare cases of autosomal recessive inheritance have also been reported with milder phenotypes in variant homozygotes and asymptomatic heterozygotes [33]. This may reflect largely functional GLUT1 transporters that tolerate the heterozygote variant scenario, but homozygosity mimics fully penetrant heterozygous missense mutations.

Synonymous variants are often assigned a non-pathogenic status with negligible impact on molecular functionality [34]. This perception is changing as biomedical investigations of synonymous and deep intronic variants and their effect on splicing may contribute substantially to the diagnostic yield for patients with rare diseases. Recent studies have challenged this notion as more sensitivity is used to detect mosaic pathogenicity and re-evaluation of NGS datasets awaits [35]. In our example, reanalysis of a recurrent *SLC2A1* variant predicted three activated cryptic splicing donor sites both upstream and downstream of the variant and confirmed by *in vitro* minigene assays. Three GLUT1-DS families now have a genetic explanation, and clinical custodians can account for the heterogenous phenotypic presentations through mosaic expression dynamics.

## CONCLUSION

This study investigated the nuanced relationship between a synonymous variant and splicing aberrations in the context of GLUT1-DS, focusing on the *SLC2A1* c.972G>A (p.Ser324=) variant. Despite the initial lack of observable splicing aberrancies, *in silico* analyses and *in vitro* minigene assays uncovered a leaky splicing defect resulting in GLUT1 frameshift truncations. This study broadens our understanding of the impact of splicing dysfunction on genetic disorders and in three families with heterogenous GLUT1-DS and epilepsy. These findings highlight the need to re-evaluate the functional consequences of synonymous variants, challenging the notion of their minimal impact in disease pathogenicity. Overall, this research contributes valuable insights into the intricate molecular mechanisms of GLUT1-DS and advocates for a more detailed approach to understanding the precise role of synonymous variants in disease pathogenesis.

## Data Availability

All data produced in the present work are contained in the manuscript

## Supplementary Information

The online version contains supplementary information with supplemental information on phenotypic presentation, 2 figures and 1 table.

## Author Contributions

MIR and SKC were involved in the conception of the study. ATH, FW and WOP were involved in generating and analysing research data. WOP, AJJ, JN, KH, JtWN and PES were responsible for the patient care and sourcing of family research materials and anonymised clinical data. SKC designed and supervised the study. ATH, FW and SKC wrote the first draft with additional contribution from WOP, MC and MIR. All authors were involved in editing, critically revising and approval of the final manuscript for submission.

## Data or materials availability

Source data and materials for this study can be made available upon request from the corresponding author. Expression and genetic databases will be updated with the pathogenic evidence presented in this study.

## Declarations

### Ethical Approval and Informed Consent

The GLUT1-DS family in this study was consented into the WERN family study (23/WA/0002) with ethical approval provided by the Wales-REC. The proband has consented to the publication of this manuscript on behalf of the members of the family.

## Acknowledgments

We thank the GLUT1-DS family for their consent, courage, and participation in our ongoing study of familial epilepsy. We would like to acknowledge Glasgow’s Children Hospital for their commissioned gene-panel findings.

