## Supplementary Information for "A synonymous *SLC2A1* variant causes familial epilepsy and paroxysmal exercise-induced dyskinesia by creating aberrant mosaic splicing patterns"

### Supplementary Material

#### GLUT1-DS Family Phenotype

The family has a complex GLUT1-DS phenotype, with family members experiencing early-onset absence epilepsy, focal (likely occipital-onset) epilepsy, paroxysmal exercise-induced dyskinesia, and intellectual difficulties.

**III-1** has had focal seizures since infancy. Her seizures consist of seeing flashing lights and blobs with occasional focal to bilateral convulsive seizures. She has had episodes of leg stiffening after exercise which have been relieved by rest or food.

The proband (**IV-2**) had episodes of falls with retained consciousness, as a young child, which were provoked by exercise or lack of food. These episodes persisted infrequently as episodes of leg cramping and twitching with abnormal dystonic posturing. She had her first epileptic seizure in late childhood and continues to get focal seizures with altered awareness (seeing coloured bubbles) and focal to bilateral convulsive seizures. One EEG showed focal left slowing, other EEGs and MRIs have been normal.

**IV-4** had absences and generalised tonic-clonic seizures starting in young childhood and persisting throughout adulthood. EEGs have shown generalised spike and wave discharges, CSF:plasma glucose ratio=0.52.

**IV-3** has epilepsy with convulsive seizures with occasional episodes of visual disturbance. EEGs have been normal

**V-1** had an unprovoked focal to bilateral convulsive seizure as an infant. Since then focal seizures with loss of awareness (seeing coloured blobs and shapes with difficult to describe feeling) and focal to bilateral convulsive seizures have continued. V-1 also has episodes of leg dystonia after exercise and a single febrile seizure. There are no developmental problems and EEGs have been normal.

**V-2** started having seizures shortly after birth. Seizures, both non-motor and convulsive generalised seizures have continued every few months. V-2 also has several episodes of limb dystonia and severe intellectual disability. V-2 is the only individual in the family with confirmed intellectual disability. Cerebrospinal fluid (CSF):blood glucose ratio was 0.42, MRI brain was normal and an EEG showed a few bursts of interictal generalised spike and wave.

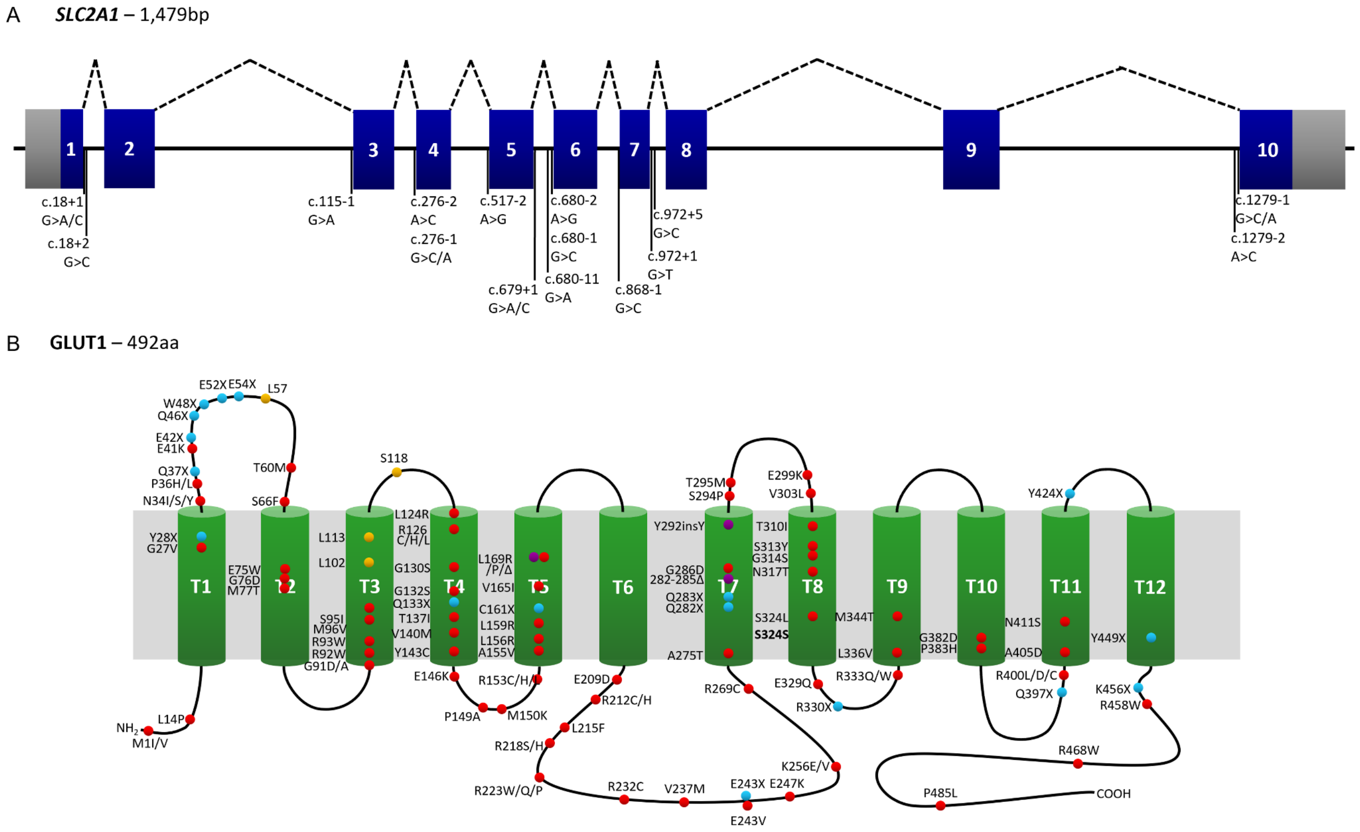


**Supplemental figure 1.** The schematic of *SLC2A1* and GLUT1 with GLUT1-DS variants shown. A) Isoform 1 comprises of exons 1 to 10. Splice-altering published variants in GLUT1 DS are shown. B) The 492 amino acid GLUT1 protein has 12 transmembrane domains. The published GLUT1 variants in GLUT1-DS are shown (red=missense, blue=nonsense, purple=in-frame indels, yellow=frameshift). The p.Ser324= (S324S) variant analysed in this study is bolded. T=transmembrane domain

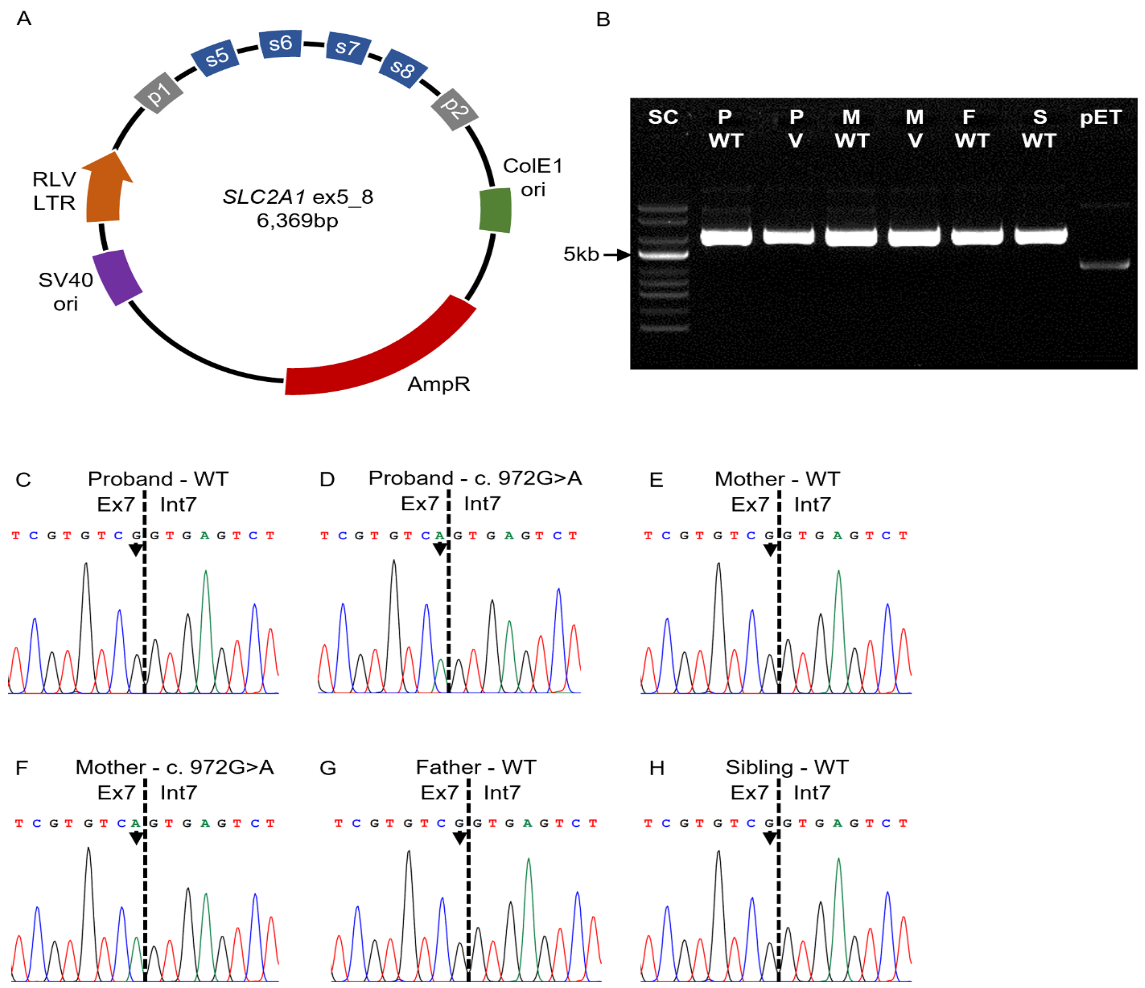


**Supplemental figure 2.** The minigene plasmids generated for the *SLC2A1* splicing assay. A) A schematic of the pET01-SLC2A1-ex5_8 minigene plasmids. Vector exons (p1 and p2, grey) flank the *SLC2A1* genomic fragment containing exons 5 to 8 (blue). Rous sarcoma virus long terminal repeat promoter (RTV LTR, orange). *ColE1* origin of replication (green). SV40 origin of replication (purple). Ampicillin resistance gene (red). B) An agarose gel showing the minigene plasmids generated for proband (P), mother (M), father (F) and sibling (S). 5kb band on the ladder is arrowed. SC = supercoiled ladder, WT = wild-type, V = variant. C-H) Sanger sequence chromatograms showing the variant status of the minigene plasmids. The variant position is arrow.

**Supplemental Table 1.** A summary of the bioinformatic tools used to predict splicing outcomes.

| Splice Program | URL | Input | Calculation method | Output | Reference | |
|---|---|---|---|---|---|---|
| HSF 3.1 | http://www.umd.be/HSF3/ | Ensembl ENST# & cDNA# with variant | Position Weight Matrices (0 to 100), MaxENT (-20 to +20) | Changes to WT & cryptic splice site strength, branch point, & enhancers/silencers (broken or gained) | Desmet *et al.*, 2009 | [1] |
| NetGene2 Server | http://www.cbs.dtu.dk/services/NetGene2/ | 200-100,000 nucleotides (WT & variant) | Confidence (0-1) | Predicts DS & AS (20 nucleotides per site), trained by a human exon/intron dataset | Hebsgaard *et al.*, 1996 | [2] |
| NNSPLICE 0.9 | http://www.fruitfly.org/seq_tools/splice.html | 100-200,000 nucleotides (WT & variant) | Score (0-1) | Scores potential DS & AS (15 nucleotides for DS, 41 nucleotides for AS), trained by human & *D.melanogaster* exon/intron datasets | Reese *et al.*, 1996 | [3] |
| FSPLICE | http://www.softberry.com/berry.phtml?topic=fsplice&group=programs &subgroup=gfind | >100 nucleotides (WT & variant) | Weight (0-16.5) | Weights (strength) of potential donor and acceptor sites (12 nucleotides per site) | Salamov & Solovyev, 2000 | [4] |
| MaxENTScan (5' SS) | http://genes.mit.edu/burgelab/maxent/Xmaxentscan_scoreseq.html | 9 nucleotides (3exon/6intron) (WT & variant) | MaxENT (-20 to +20), MDDM, FMM, WMM | Scores a specific DS from 4 algorithms based on nucleotide sequence | Yeo & Burge, 2004 | [5] |
| SROOGLE | http://sroogle.tau.ac.il/ | Upstream intronic, exonic, downstream intronic nucleotide sequence (user defined) | Donor: PSSM (0-100), ΔG Acceptor: PSSM (0-100) | Weights (strength) of potential DS, AS, branch sites & PP tracts. Shows enhancers/silencers from 13 datasets. | Schwartz *et al.*, 2009 | [6] |
| CRYP-SKIP | http://cryp-skip.img.cas.cz/ | 100bp upstream intronic, exonic, 100bp downstream intronic nucleotide sequence (user defined) | EXSK (0-0.5) vs CR-E (0.5-1), CS (0-1) | Probability of exon skipping (EXSK) vs cryptic splice site activation (CR-E). Confidence of cryptic splice site utilisation | Divina *et al.*, 2009 | [7] |
